# Toward more patient-centered diabetes care in Switzerland: Patient perspectives and practical solutions

**DOI:** 10.64898/2025.12.06.25341746

**Authors:** Odile-Florence Giger

## Abstract

This study investigates the challenges and enablers of self-management in type 2 diabetes, the role of primary care in diabetes management, and patients’ preferences for additional support services in Switzerland. Participants reported significant difficulties in sustaining behavioral changes and emphasized the need for more structured guidance to improve self-management. Many felt unsupported in primary care, stating that their general practitioners could have intervened earlier when they were at high risk but before an official diagnosis. Additionally, patients highlighted the lack of centralized, up-to-date diabetes resources in Switzerland, contrasting it with structured systems like the National Health Service in the United Kingdom. There was also a strong demand for a unified digital health platform to consolidate glucose readings, blood pressure data, and medication lists, ensuring seamless integration with healthcare providers. These findings underscore the need for patient-centered diabetes management strategies that integrate digital innovations, proactive primary care interventions, and accessible, evidence-based resources to enhance self-management.

## Introduction

Globally, chronic diseases account for 49% of the total disease burden and 60% of deaths.^1^ In high-income countries, they represent the leading cause of disability-adjusted life years and are projected to become the primary cause of death by 2030.^2^ Diabetes - characterized by both micro- and macrovascular complications, as well as reduced health status and quality of life - poses a significant public health concern worldwide.^3^ In high-income countries, it ranks as the eighth leading cause of disability and mortality, and its prevalence is predicted to continue rising in the coming decades.^4^

In Switzerland, where approximately 6-7% of adults are estimated to have Type 2 Diabetes (T2D), it is perceived as a growing challenge due to the country’s aging population and changing lifestyle factors.^5,6^

Managing T2D requires continuous self-management and behavioral changes,^7^ yet many patients struggle with adherence to treatment guidelines.^8^ Effective management involves maintaining a healthy diet, regular physical activity, medication adherence, and routine monitoring of key health indicators such as blood glucose levels.^9^ However, many patients face challenges in integrating these behaviors into their daily lives, leading to suboptimal disease control. ^8^ Research shows that while patients perceive themselves as well-informed, many still lack a full understanding of crucial health markers like HbA1c levels. ^10^ Furthermore, long-term behavioral change is difficult, and many individuals require ongoing support to sustain healthy habits.^7^

Beyond self-management, primary care plays a central role in diabetes treatment, as general practitioners (GPs) are often the first point of contact for patients treating over 90% of the individuals with T2D.^11^ Many rely on their GP for routine check-ups, medical advice, and coordination of care. ^11^ However, studies indicate that only 44% of T2D patients treated by GPs reach their target HbA1c level, ^12^ highlighting gaps in primary care support. Challenges such as high GP workloads ^13^, limited consultation time, and inconsistent adherence to diabetes guidelines can hinder effective management. ^14,15^

Given these gaps in self-management and primary care, there is increasing interest in additional external support that could enhance diabetes management. Digital health tools, coaching programs, and community-based initiatives have shown potential benefits,^16,17^ yet it remains unclear what types of support patients would find most useful.^18^ Furthermore, many of these services are not covered by health insurance, requiring out-of-pocket payments from patients.^19^ While diabetes management has been widely studied in various healthcare systems, there is limited research focusing on patient preferences and gaps in diabetes care within Switzerland. Understanding these perspectives is critical for developing patient-centered interventions and optimizing diabetes management strategies. To address these gaps, this study explores from a patients perspective how T2D management can be improved in Switzerland by examining three key areas:

1. **Self-Management & Behavioral Change:** What are patients’ main challenges in managing T2D, and what enables them to improve self-management?
2. **Primary Care:** What challenges do patients face in primary care, and how can GPs better support patients in diabetes management?
3. **Additional Support & Willingness to Pay:** What additional support do patients desire, and what services would they be willing to pay for?

To answer these research questions, twelve semi-structured interviews with T2D patients residing in Switzerland were conducted. The interviews aimed to capture in-depth insights into their experiences, perceptions, and needs.^20–22^ This qualitative method allows for open-ended discussion, enabling participants to share personal stories and reflect on how T2D could be optimized. The interviews were audio-recorded, transcribed verbatim, and analyzed thematically to identify common themes, key challenges, and potential areas for intervention.

By focusing on this underexplored context, the study offers valuable insights into the Swiss healthcare setting and lays the groundwork for new innovations.

## Theoretical Background

To understand the patients perspective better the relationship between patients and healthcare providers needs to be examined. Information imbalances can lead to poor choices, affecting treatment adherence and health outcomes.^23^ The Principal-Agent Theory helps explain these issues by highlighting misaligned incentives and communication gaps^23^ and provides insight into structural barriers and potential solutions for improving patient-provider interactions.

The principal-agent theory is a fundamental concept in economics and organizational studies that describes the relationship between two parties: a principal, who delegates authority, and an agent, who acts on behalf of the principal.^23^ The problem deteriorates when there is a greater discrepancy of interests and information between the principal and agent, as well as when the principal lacks the means to punish the agent.^24^ The core issue within this relationship is information asymmetry - where the agent typically has more knowledge and expertise than the principal - leading to potential conflicts of interest.^23^ Agents may not always act in the best interest of principals due to differing incentives, leading to agency costs, such as inefficiencies, moral hazard, or adverse selection.^25^

In healthcare, physicians act as agents, while patients serve as principals. This relationship is characterized by asymmetry in knowledge:^26^

- Physicians possess superior medical expertise, including knowledge of diseases, available treatments, and potential risks.^26^
- Patients, on the other hand, have better insight into their own lifestyle preferences, treatment adherence tendencies, and beliefs about illness and medication.^26^

This asymmetry can result in suboptimal healthcare decisions. Especially in fee-for-service models, physicians may be incentivized to increase service volume (e.g., overprescribing medications or recommending unnecessary treatments) to maximize revenue, rather than strictly prioritizing patient well-being.^27^ Also, time constraints from the physicians side may limit shared decision-making, resulting in rushed consultations and inadequate patient education.^26^

While Principal-Agent theory traditionally positions the physician as the agent acting on behalf of the patient, the reverse dynamic is especially relevant in the context of T2D management.^28^ In this case, the patient functions as the agent, as they are responsible for implementing daily self-care tasks such as blood glucose monitoring, medication adherence, physical activity, and dietary choices. The GP becomes the principal, relying on the patient’s accurate self-reporting and consistent behavior to make informed medical decisions.^28^ However, when patients fail to disclose challenges such as difficulties with diet, emotional stress, or non-adherence to medication or delay seeking help, this creates a form of information asymmetry that limits the GP’s ability to intervene early or adjust treatment plans effectively.^29,30^

For example, a study found that 25.3% withheld information about unhealthy diets, while 23.5% did not disclose their failure to exercise regularly.^29^ Patients cited fear of judgment and a desire to appear cooperative as primary reasons.^29^ This reversal illustrates how agency problems can emerge from the patient side as well and underscores the importance of mutual trust and open communication in chronic disease management.

## Methods

The primary data collection approach involved conducting qualitative semi-structured interviews with T2D patients.^31^ Qualitative interviews reveal the complexity of individuals’ experiences, including their thoughts, opinions, and perspectives.^20–22^

This study adhered to the reporting guidelines established by the Consolidated Criteria for Reporting Qualitative Research (COREQ)^32^

This study was conducted in accordance with the Declaration of Helsinki. This study did not involve the collection of clinical or otherwise sensitive health data. In view of its minimal-risk profile and its exclusive focus on expert knowledge and lived experience, the study was exempt from a formal review and approval by the Ethics Committee of the University of St. Gallen. Participants were fully briefed on the study’s purpose and procedures before the interviews. Written consent was obtained, including permission to record audio. Participants were informed of their right to withdraw at any time, and all data were anonymized and securely stored to ensure confidentiality.

### Development of the interview guide

The author adopted a stepwise process to develop the interview guide. First, the author identified the prerequisites for using semi-structured interviews to shape the guide. The author then conducted a literature review on patients’ perceptions of the current Swiss healthcare system. Next, the author designed a preliminary semi-structured interview guide comprising two tiers: main themes and follow-up questions. Every participant was queried on the main themes, and both pre-designed and spontaneous follow-up questions were used to encourage deeper exploration of specific topics.^33^ Throughout the interview phase, the author iteratively refined the questions multiple times as new insights emerged.

### Participant recruitment and selection process

The author recruited the participants through Testing Time, an online participant recruitment website that pay interviewees for participating. All the participants spoke German or English and lived in Switzerland and had to be diagnosed with T2D. Recruitment concluded once theoretical saturation was achieved, defined as the point at which three consecutive interviews yielded no additional concepts.

### Data collection and data analysis

The author conducted twelve in-depth interviews between January 2025 and March 2025 with individuals with T2D. Each interview was conducted in an open-ended, semi-structured manner, providing participants with the opportunity to discuss significant topics and allowing researchers to probe more deeply where clarification was needed.^34^ Each interview began with participants offering personal introductions, followed by an overview of the study’s framework (see Appendix B.1 for the interview guideline).

All interviews were recorded digitally using Microsoft Teams, generating 142 pages of transcripts. Each transcript was thoroughly reviewed for accuracy, anonymized, and assigned a unique study ID. Data collection and analysis proceeded simultaneously using ATLAS.ti software.

The interview process itself was divided into two phases. In the first phase, participants were asked about their sociodemographic background, followed by an exploration of the daily challenges in managing T2D and potential improvements they wish for. In the second phase, the discussion focused on external support possibilities and willingness to pay. At the conclusion of each interview, participants were invited to pose any additional questions. The data was analyzed inductively to allow themes to emerge from the participants’ experiences. Figure 1 illustrates the coding framework derived from the qualitative interviews, structured according to the study’s three main research questions. The three central categories—Self-Management & Behavioral Change, Primary Care, and External Support & Willingness-to-pay—are further subdivided into specific themes and subthemes identified through inductive analysis. This categorization allows clear visualization of patient challenges, enabling factors, desired improvements in primary care, and preferred additional support services.

**Figure 1.**
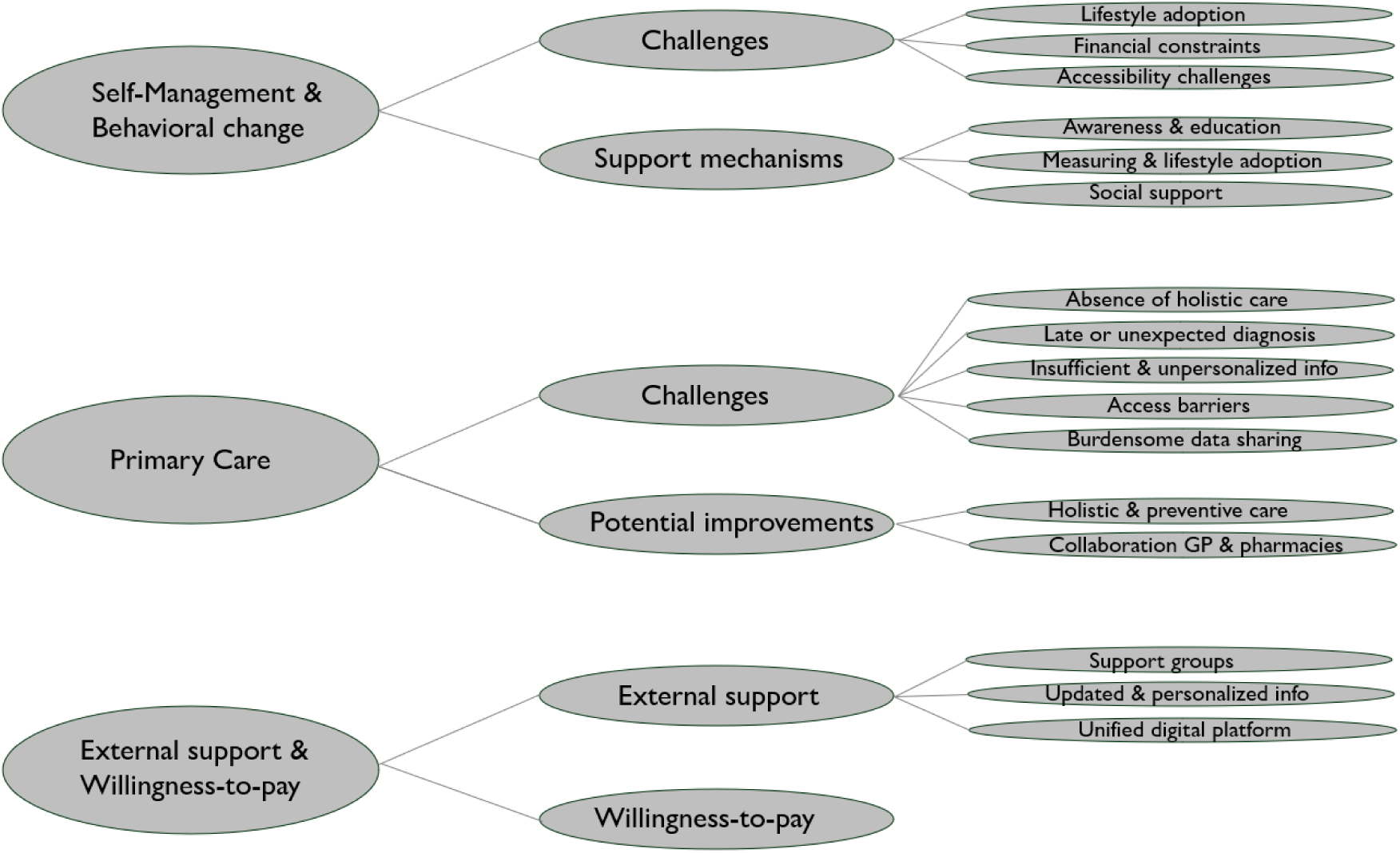
Coding of main themes into the predefined categories “Self-Management & Behavioral change”, “Primary Care”, “External support & Willingness-to-pay”

## Results

### Self-Management & Behaviral Change Challenges

Interviewed patients reported significant challenges in maintaining consistent self-management routines, citing multiple barriers related to **lifestyle adoption, financial constraints, and accessibility challenges.**

- From a **lifestyle adoption** perspective, a recurring theme was difficulty maintaining dietary discipline, particularly among individuals living alone. Several participants indicated that they rarely cooked for themselves, leading to a reliance on convenience foods that often lacked the nutritional quality necessary for optimal diabetes management. Interviewee 2 noted, *“I know it might be better if I cook more by myself but it’s hard if you have to do that all the time being just alone.”* Another prominent challenge was low motivation for physical activity, often exacerbated by joint pain. Many participants expressed frustration over their inability to engage in regular exercise, despite recognizing its importance. Interviewee 6 stated, *“I would like to move more, but my knees hurt too much, and that makes it really difficult to stay active.”* Furthermore, one patient shared that receiving the diagnosis was such a profound shock that she struggled to accept it for several years. During this period of denial, she found it difficult to engage with the necessary lifestyle changes. Her experience highlights how the emotional impact of a chronic illness diagnosis can act as a barrier to early self-management and delay the adoption of healthier behaviors.
- **Financial constraints** further complicated diabetes self-management, particularly in relation to the cost of blood glucose test strips. Several participants highlighted that regular monitoring was financially burdensome, limiting their ability to track blood sugar levels as frequently as recommended. Interviewee 2 shared, *“One test strip costs almost 1 frank. If you want to test regularly it gets really expensive.”*
- **Accessibility challenges** also emerged as a key barrier. Some participants reported that they previously relied on structured exercise programs, such as Aqua Gym, but struggled to find alternatives after these programs were discontinued. Others indicated that the distance to fitness centers was a deterrent, with one stating, *“The gym is too far I would need to take the car to get into another village and that’s an extra effort so I rather stay at home and go for walks sometimes.”*(Interviewee 12)

### Support mechanisms

Interview participants highlighted three key themes influencing their ability to manage the condition effectively: **Awareness and education, measuring and lifestyle adaptation, and social support.**

- In the field of **awareness and education** many participants emphasized that it helped them to see their parents suffered from diabetes and also getting informed about the severe long-term consequences when not managing it well. (e.g. foot amputation) This motivated patients to try to improve their diets or move more. This was mentioned by interviewee 7: *“A major incentive for me today is my polyneuropathy in my feet, which is sometimes painful. Since I know that diabetics sometimes have to have a toe or even a foot amputated, I naturally do everything I can to prevent that as much as possible.”* And interviewee 1 mentioned that her previous heart attack has given her more awareness to take better care over her weight: *“I’m sure I also lost weight because I was very motivated after being ill, specifically with my heart. That certainly supported the weight loss.”*
- Also regular **measuring and lifestyle adaptation** helped patients. Some participants described how frequent blood sugar measurements helped them remain disciplined in their dietary choices. Interviewee 2 shared their personal strategy: *“I measure my blood sugar myself every other morning, which keeps me motivated: in the evening, I know that I will be extremely upset in the morning if my blood sugar is over 8 again. If it is below that, it reassures me and helps me stay focused on my goals. If my blood sugar is repeatedly well above 8 in the morning, all alarm bells go off, and I immediately start tightening control over my eating habits.”* Beyond blood sugar monitoring, regular exercise was seen as beneficial to control the blood sugar levels. This was stated by interviewee 10: *“The weekly use of my home bike helps, even if it’s always just 10 km that I ride!”*
- Next, **social support** was crucial for maintaining long-term behavioral changes. Some participants mentioned the positive impact of partners assisting with dietary changes. Others found motivation through structured online programs and support groups, which provided encouragement and shared knowledge on managing the disease effectively. Interviewee 8 mentioned: *“So like I said when I was researching if there are support groups, I found groups in the US which are medically backed. This helped me a lot.”*

### Primary care

#### Challenges

- A prominent theme was the perceived **absence of holistic care,** as many participants felt that GPs addressed only one problem at a time not coordinating with other doctors. Also, participants described time constraints during consultations and a lack of integrated or personalized treatment. This was mentioned by interviewee 8: *“She then finally put me on to a psychiatrist because she realized that I needed help also with managing my mental struggles. So, you know, like with pregnancy, for example, the gynecologist is there, but they don’t really have knowledge of diabetes, so they say you have to work with an endocrinologist or GP and they all don’t talk to each other, so everything is an isolated system.”* They also reported that their concerns outside a single, immediate issue often went unanswered and that the onus was on patients to manage and coordinate additional care on their own such as for example psyichaitrst. This was mentioned by interviewee 8: “*Again, I mean, she referred me to a psychiatrist, but I had to go and find my own.”*
- A recurring theme among participants was the **late or unexpected diagnosis** of diabetes. In many accounts, individuals learned of their condition by chance—during unrelated medical procedures, gynecological check-ups, or emergency hospital admissions. This was mentioned by interviewee 6: *“I wanted to do a hair transplant, so they had to take blood. And then suddenly the doctor mentioned that I have type 2 diabetes, although I was in frequent care with my GP.”* Others had been in a “borderline” range across several consultations, yet only received a formal diagnosis after persistently requesting further blood glucose measurements, often because of personal health concerns or family history. Participants felt that GPs could have intervened earlier, warning them of diabetes’ potential severity and outlining preventive measures rather than waiting for definitive indicators before providing treatment guidelines. This was mentioned by interviewee 2: *“My GP never told me to change my lifestyle habits although I was always at the upper limit. Only from the point when diabetes actually broke out he intervened. Looking back, I actually think it’s unfortunate—I could have taken better care of myself earlier, if I had known the consequences.”*
- Many participants reported receiving **insufficient and impersonalized information** regarding diabetes management, which hindered their ability to grasp the full severity of their condition. Although an abundance of material was available online, individuals noted discrepancies and inconsistencies that made self-education challenging. Also some participants mentioned that they get most of the information from patient flyer or from health magazines but this is often very outdated generic information. This was mentioned by interviewee 8 in the following way: *“Some doctors had their education many years ago, so 10, 20, 30 years ago. And obviously research has found many new insights, but if they don’t know, they don’t know it.”* Several participants also felt that their GPs failed to offer clear answers, leaving many questions unanswered or deterring some from seeking clarification. In particular, dietary guidance from nutritionists was perceived as not personalized enough to the own needs, prompting frustration among those requiring specific modifications—for instance, advice that did not account for dietary restrictions such as gluten or dairy intolerance. This lack of individualized, actionable advice ultimately undermined the perceived usefulness of nutrition counseling and exacerbated feelings of confusion and uncertainty. Interviewee 5 stated the following: *“But then I finally went to the nutritionist and I was completely disappointed. If you have diabetes, that’s one thing, but I also have grain intolerance and a dairy intolerance. I’m not allowed to eat any dairy at all, not even camel milk—just nothing. And I really noticed that the nutritionist was desperate. She couldn’t apply any standard approach. In the end, I was the one advising her because she had never dealt with such a complex case before. Although, I’m convinced there exist many more patients with these intolerancies.”* Furthermore, also no information about new diabetes devices was shared. One third of the participants either have never heard of a continuous glucose monitor or they did not know what it was for. Interviewee 12 mentioned: *“I have never heard about this continuous glucose monitor. I don’t really get such information from my GP, although probably there exist already a lot of innovation in diabetes management.”*
- Furthermore, **access barriers** such as extended waiting times and limited clinic hours created significant barriers for participants seeking appointments or prescription refills. When it comes to waiting times, some reported wait times of up to three months, with staff appearing unresponsive to urgent needs. This was stated by interviewee 8 in the following way: *“If you need something urgently, then the minimum wait is three months. The receptionist is handling your appointment call, so they don’t have the sense that how to prioritize or triage cases.”* Limited clinic hours were another barrierer. Early morning fasting tests often conflicted with strict work schedules, requiring time off or frequent rescheduling. These factors, compounded by family responsibilities, discouraged many from attending quarterly check-ups—some had to use vacation days just to make an appointment. As a result, critical follow-ups and medication adjustments were delayed, complicating diabetes management. This was mentioned by interviewee 4: *“I can’t make it to go every 3 months to the doctor to do the blood sugar test again. I have to go on an empty stomach, which means in the morning. But I work every morning, so from that perspective, it’s not even possible. I always have to take vacation days for the GP visit or find another way to be able to go. That’s why I’m going only 1-2x per year.”*
- Next, **burdensome data sharing** with the GP posed another challenge. Participants often relied on multiple devices to measure and track different health parameters—ranging from blood pressure to blood glucose levels. However in most of the cases sharing the health data with their GP was considered as burdensome. Especially, consolidating these data points into a single, comprehensible format proved time-consuming. Some resorted to printing out PDF files or manually writing their readings on paper before sharing them with their GP. This fragmented approach not only added extra effort but also limited the ability to monitor trends over time, ultimately impeding more effective diabetes management. This was mentioned by interviewee 1: *“So I print the blood pressure data from the app and bring it to the doctor. It’s just extra effort that is tedious, but I want the doctor to have it as a document in front of them. Because most of the time, feel the doctor doesn’t even want to know what my values are that I measured at home in my app.”*

### Potential improvements for primary care

- Participants strongly emphasized the need for a more **holistic care together with an early screening program**. Rather than focusing solely on isolated metrics, they felt that GPs should attend to the patient’s overall well-being, (eg. ask about their private life and their psychological well-being) and proactively identify risk factors before a formal diagnosis occurs. In particular, individuals who had borderline test results for an extended period would have wished for earlier warnings or guidance on the potential severity of diabetes. Early screening, particularly for individuals with obesity, was highlighted as crucial. A more comprehensive, patient-centered approach could enable proactive interventions, improve long-term health outcomes, and cultivate a stronger sense of collaboration between patients and healthcare providers. This was mentioned by interviewee 11: *“I guess if I had more information about diabetes type 2 from my doctor or health insurance I would have taken more consideration about that illness. If my doctor would have informed me about the actions I could take to help my illness instead of given me pills without informing me what I could do to help me overcome the illness.”* and interviewee 6 added: *“It would certainly be better if there were a more mandatory form of prevention. For overweight individuals like me, there should be incentives for blood sugar prevention.”*
- Other participants expressed a desire for closer **collaboration between GPs and local pharmacies** to accommodate busy schedules and long distances to their GP. For instance, they suggested allowing individuals to measure their HbA1c levels at conveniently located pharmacies—such as during early morning hours before work—and then discuss the results over the phone with their GP. This approach would simplify the diagnostic process, improve access to timely treatment, and make it easier to schedule and attend regular check-up appointments, for example every three months. This was mentioned by interviewee 4: *“It would be helpful if I just could test my blood at the pharmacy and the pharmacy would send the results to my GP so that we could discuss this over the phone if there are adjustments needed.”*

### External support and willingness-to-pay

#### External support

- Some participants mentioned that **support groups** could help them to stay motivated and up to date with the most important information. However not all participants were a fan of support groups. Interviewee 8 said: *“Support groups would help me to stay motivated and to get additional tips from others”.* While interviewee 10 mentioned: *“I’m rather the “lonely wolf”-type of person, I don’t like social interaction and groups so much.”*
- Participants stressed the importance of having reliable, **up-to-date, and personalized information** sources to guide their diabetes management. A proposed solution involved developing dedicated platforms or magazines that offer trustworthy, regularly scientifically updated content, specifically tailored to individual needs and health profiles. This was mentioned by interviewee 8: *“Like, if you look at the UK, the NHS has a separate department for diabetes, where they really put out a lot of information very frequently and they’re updating their knowledge. They have conferences and all of that, but Switzerland is very, very siloed. For example you can also look up the coaching plattfrom that I use:* Mastering-Diabetes.org*, nutrition* facts.org*. These are very helpful resources that don’t exist in Switzerland.”*
- Many participants expressed a strong desire for a **unified digital platform** that consolidates all relevant health data—ranging from blood glucose readings and blood pressure measurements to medication lists—in one place. Ideally, this system would connect directly with the GP’s records, eliminating the need for manual data entry or repeated updates. In addition, patients hoped to see their health trends over time, enabling them to observe patterns in their readings and better understand the progression of their condition. Interviewee 6 mentioned this in the following way: *“I’m right now setting up the electronic health record. Ideally I could submit all my data from the blood pressure monitoring app to one single platform and have an overview.”*

#### Willingness-to-pay

- When asked how much they would personally pay, in addition to their health insurance costs, for a solution that could help them achieve normal blood glucose levels, participants gave mixed answers. More than half of the participants mentioned that they would pay between 1800 – 2400 Swiss franks per year. This was mentioned by interviewee 2: “*For a convincing and highly promising measure, I would certainly be willing to contribute to the costs, for example, around 30% of the costs, up to an annual amount of CHF 2,000.”* Other participants could not afford to pay extra or expected the health insurance to pay for them. For example interviewee 12 stated: *“Since I live at the subsistence level, I can hardly afford any additional expenses. The only thing I could possibly imagine is canceling my TV subscription to save a little money—maybe 10 francs per month. Exercise is important, but I can’t spend anything on courses or gyms either.”*

## Discussion

The findings from this study highlight the multifaceted challenges that patients with T2D face in self-management and primary care. These challenges align with existing literature on diabetes care, which emphasizes the importance of behavioral change, structural support, and proactive primary care interventions.^35^

### Self-Management and Behavioral Change: Challenges and support mechanisms

Our findings corroborate previous research indicating that maintaining consistent self-management routines is particularly difficult for individuals living alone.^36^ A notable proportion of participants expressed difficulty in adhering to dietary recommendations, with many relying on convenience foods due to the burden of cooking for one person. Swiss national data indicates that over 38.4% of adults live alone, a demographic trend that may exacerbate unhealthy eating patterns.^37^ Also, on the other side previous literature shows that patients achieve a lower HbA1c level if they are married, not living alone. ^38^ Furthermore, limited motivation for physical activity, often due to joint pain, was a recurring theme. This is consistent with estimates suggesting that more than 50% of individuals with T2D experience some degree of musculoskeletal pain, which can significantly hinder physical activity engagement.^39^ Financial constraints emerged as a substantial barrier, particularly regarding the affordability of blood glucose test strips. This financial burden is not trivial, particularly for those on fixed incomes, and reinforces the need for insurance policies that expand reimbursement coverage for essential diabetes supplies. Despite these challenges, various support mechanisms proved instrumental in promoting behavioral change. Awareness of diabetes complications, such as foot amputation or cardiovascular risks, emerged as a strong motivational driver. This finding is supported by prior research suggesting that individuals who perceive severe health risks are more likely to engage in preventive health behaviors.^40^ Nevertheless, this is not always the case: in adolescents with high body mass indey the risk perception alone is not sufficient to sustin healthy behaviors.^41^ Similarly, regular blood sugar monitoring played a pivotal role in reinforcing dietary discipline, consistent with studies demonstrating that self-monitoring is associated with better glycemic control and improved long-term adherence.^42^

### Primary Care: Gaps and opportunities for improvement

A key concern expressed by participants was the lack of holistic and coordinated care among GPs and specialists. Many reported feeling that healthcare professionals addressed only isolated symptoms rather than adopting a comprehensive approach to diabetes management. This reflects a principal-agent problem in which healthcare providers (agents) operate under different incentives than the system (principal). Fee-for-service models often prioritize reactive treatment over preventive, long-term care, leading to fragmented patient experiences.^26^

Delayed or incidental diabetes diagnoses were another common theme, with several participants reporting that their condition was only identified during unrelated medical procedures. This is concerning, given that early detection is crucial for preventing complications. Most recent data indicates that slightly more than one-quarter of adults with diabetes had undiagnosed diabetes.^43^ This underscores an information asymmetry issue between patients (agents) and healthcare providers (principals). Without systematic screening mechanisms, patients remain unaware of their condition, and providers may miss early intervention opportunities. In this context, inexpensive early screening programs, for example administered by specialized diabetes nurses, may represent a cost-effective approach to facilitate screening and reduce the risk of long-term complications. Nevertheless, some patients reported feelings of shame related to their diabetes, which led them to avoid seeking information and resist accepting their diagnosis. This emotional response may result in patients withholding important information from their GP—illustrating a reversal of the typical principal-agent dynamic, where the patient assumes the role of the agent and the GP the role of the principal.

Another critical issue was the lack of up-to-date, personalized information on diabetes management. Several participants noted discrepancies in available resources and received conflicting advice from healthcare providers. This study found that one third of the participants had never heard of continuous glucose monitors, despite strong evidence supporting their benefits in glycemic control for T2D patients.^44^ This imbalance of information causes patients to make less effective decisions about their health, as they lack the necessary knowledge or guidance. From a principal-agent perspective, this reflects a breakdown in communication and incentives between physicians and patients, making it harder for patients to act in their best interest.^26^ This information gap highlights the need for improved patient education initiatives and a greater focus on digital health literacy. Notably, in the U.S., the monthly rate of continuous glucose monitor prescriptions for new users increased by 36% from 2020 to 2021, with a 125% rise in primary care settings.^45^ Whether this trend will be replicated in Switzerland remains uncertain.

A key recommendation from participants was a shift towards a more preventive and patient-centered care model. Proactive risk identification and lifestyle counseling before diabetes onset could mitigate long-term complications. This aligns with international best practices, where structured lifestyle interventions have been shown to reduce diabetes incidence by up to 58% in high-risk individuals.^46^ However, the principal-agent lens suggests that a lack of direct financial incentives for GPs to focus on prevention contributes to the existing gap in early intervention.^47^

Additionally, the findings suggest a strong need for the implementation of a low-cost, scalable screening solution for T2D in Switzerland. Such a screening approach could facilitate earlier diagnosis and intervention, addressing a key challenge highlighted by participants regarding late or unexpected diagnoses. A cost-effective, widely accessible screening program—potentially utilizing local pharmacies, or specialized diabetes nurses —would not only enable proactive preventive care but also significantly reduce long-term healthcare expenditures and complications associated with delayed diabetes management. Several participants suggested that HbA1c testing at pharmacies, with results digitally transmitted to GPs, would enhance accessibility. In the UK the Company Chemists Association has advocated for national diabetes screening within local pharmacies. Their report suggests that such a service could screen up to 1.5 million adults annually, identifying approximately 45,000 individuals with undiagnosed diabetes and 180,000 with pre-diabetes each year.^48^

### Additional support and willingness-to-pay

Many participants expressed a strong interest in integrated digital platforms for consolidating health data and facilitating communication with GPs. Currently, the fragmentation of health data requires patients to manually transfer readings, a cumbersome process that hinders trend monitoring. For example, the digital health platform Glooko showed that patients using their health platform could decrease their Hba1c level by 12%.^49^ Nevertheless, not all patients were excited about sharing health information.

Also, participants highlight the diverse needs of individuals managing diabetes, emphasizing tailored support, reliable information, and integrated digital solutions. Participants stressed the importance of centralized, up-to-date diabetes information, noting gaps in Switzerland compared to structured models like the NHS. A recent study suggests that access to health information reduces patients’ uncertainty and increases compliance.^50^ Better access to information could also lessen the information between patients and GPs.^51^ While some participants valued support groups for motivation, others preferred independent management, suggesting a need for flexible engagement options. Finally, the willingness-to-pay analysis revealed significant variation, with some participants indicating they would contribute up to CHF 2,400 per year for an effective intervention, while others could not afford additional costs. This underscores the importance of tiered pricing models or public-private partnerships to ensure equitable access to diabetes management solutions. A study in Europe has shown that T2D patients are willing to pay up to €128 per year for individual consultations and €97 per year for 10 kg of anticipated weight loss in lifestyle programs additionally to their health insurance.^52^

### Final recommendations

To sum up, the following points are final recommendations that primary care and the healthcare system should address to better support T2D patients:

### Primary care

1. **Establish a low cost scalable screening solution:** This could involve accessible blood tests at local pharmacies or labs, with results digitally shared with GPs or health coaches. Patients would then receive follow-up advice via phone or digital consultations. This low-threshold approach enables early detection and timely support while easing the burden on primary care. Furthermore, GPs should adopt a more proactive role in diabetes prevention, conducting routine screenings that could also be administered by specialized diabetes nurses.
2. **Improve care coordination:** GPs should establish better communication pathways between GPs, endocrinologists, nutritionists, and mental health professionals to ensure a more holistic approach to diabetes care.
3. **Expand structured support systems:** Healthcare providers should be able to cooperate with local pharmacies and offer both in-person and virtual support programs to cater to diverse patient preferences. **Policy makers and software providers**
4. **Increase access to affordable self-monitoring tools:** Policy interventions should focus on reducing the financial burden of glucose test strips and contoinous glucose monitors for low-income patients through subsidies or insurance coverage expansions.
5. **Expanding patient education**: Digital literacy initiatives and structured education programs can bridge knowledge gaps and reduce information asymmetry.
6. **Develop a centralized digital health platform:** A centralized digital platform integrating patient health data with GP records should be developed to enhance monitoring and decision-making while minimizing administrative burdens. Ideally, personalized health information would be included in such a solution.

### Limitations

This study provides valuable qualitative insights into patient perspectives on T2D management in Switzerland; however, several important limitations must be acknowledged. Firstly, the sample size, while adequate for achieving theoretical saturation in qualitative research, remains relatively small with only twelve participants. Such a limited number significantly restricts the generalizability and representativeness of the findings across the broader Swiss population of individuals with T2D. The experiences and viewpoints expressed by the participants may not fully capture the diversity of patient experiences, particularly across varying demographic profiles, socioeconomic statuses, or geographical regions within Switzerland.

Furthermore, the study predominantly included participants from the German-speaking region of Switzerland, with minimal representation from the French-speaking region and none from the Italian-speaking part. This geographical bias may limit the applicability of the results to regions with different healthcare infrastructures, cultural practices, or patient engagement strategies. Future research should aim for a more balanced geographical representation to enhance the robustness and generalizability of the findings.

Additionally, reliance on self-reported data introduces potential biases such as recall bias and social desirability bias. Participants may unintentionally omit or exaggerate their experiences, behaviors, or attitudes, which could skew the reported findings. Although qualitative methods excel at capturing nuanced perspectives, triangulating interview data with quantitative measurements or observational data could mitigate these biases and provide more comprehensive insights.

Another limitation pertains to the participant recruitment method via the paid platform Testing Time. This approach may have introduced selection bias, potentially favoring individuals who are more digitally literate, financially incentivized, or already engaged with research activities. Consequently, these participants may not accurately reflect the broader T2D patient population, particularly those who are less tech-savvy.

Finally, the study’s cross-sectional design limits the ability to observe changes in patient perceptions and behaviors over time. A longitudinal approach would allow for a more in-depth understanding of how self-management strategies, healthcare interactions, and patient support needs evolve throughout different stages of diabetes management

### Conclusion

This study underscores the critical need for a more patient-centered approach in T2D management, addressing the financial, logistical, and behavioral barriers that hinder self-care. Improving primary care coordination, expanding access to digital health solutions, and strengthening preventive interventions will be essential to fostering better long-term health outcomes. Future research should explore the effectiveness of integrated digital health solutions in real-world settings, assessing their impact on patient adherence and clinical outcomes. Next, a quantitative, representative study would allow for the validation and generalization of the qualitative findings presented in this paper. With a larger sample size, statistical methods could more accurately measure the prevalence and distribution of the identified challenges, preferences, and support needs across the broader Swiss population. This would not only improve external validity but also enable researchers and policymakers to understand how widespread the observed issues are among T2D patients nationwide.

Additionally, more studies are needed to evaluate the role of alternative care models, such as pharmacist-led diabetes management. Furthermore, research on the cost-effectiveness of subsidizing diabetes monitoring tools could inform policy decisions to improve accessibility. Investigating personalized education strategies tailored to individual patient profiles would also contribute to more effective diabetes management. By continuing to develop and test innovative approaches, healthcare systems can better support individuals with T2D in achieving sustainable and improved health outcomes.

## Data Availability

All data produced in the present study are available upon reasonable request to the authors

